# Machine Learning Prediction Models for Chronic Kidney Disease using National Health Insurance Claim Data in Taiwan

**DOI:** 10.1101/2020.06.25.20139147

**Authors:** Surya Krishnamurthy, KS Kapeleshh, Erik Dovgan, Mitja Luštrek, Barbara Gradišek Piletič, Kathiravan Srinivasan, Yu-Chuan Li, Anton Gradišek, Shabbir Syed-Abdul

## Abstract

**Background and Objective:** Chronic kidney disease (CKD) represent a heavy burden on the healthcare system because of the increasing number of patients, high risk of progression to end-stage renal disease, and poor prognosis of morbidity and mortality. The aim of this study is to develop a machine-learning model that uses the comorbidity and medication data, obtained from Taiwan's National Health Insurance Research Database, to forecast whether an individual will develop CKD within the next 6 or 12 months, and thus forecast the prevalence in the population.

**Methods:** A total of 18,000 people with CKD and 72,000 people without CKD diagnosis along with the past two years of medication and comorbidity data matched by propensity score were used to build a predicting model. A series of approaches were tested, including Convoluted Neural Networks (CNN). 5-fold cross-validation was used to assess the performance metrics of the algorithms.

**Results:** Both for the 6 month and 12-month models, the CNN approach performed best, with the AUROC of 0.957 and 0.954, respectively. The most prominent features in the tree-based models were identified, including diabetes mellitus, age, gout, and medications such as sulfonamides, angiotensins which had an impact on the progression of CKD.

**Conclusions:** The model proposed in this study can be a useful tool for the policy-makers helping them in predicting the trends of CKD in the population in the next 6 to 12 months. Information provided by this model can allow closely monitoring the people with risk, early detection of CKD, better allocation of resources, and patient-centric management

## 1. INTRODUCTION

Chronic Kidney Disease (CKD) is a life-long disease with an insufficient kidney function, where patients have to live with a compromised quality of life. It causes a heavy burden on the healthcare system because of the increasing number of patients, high risk of progression to end-stage renal disease (ESRD), and poor prognosis of morbidity and mortality. Asia has the highest prevalence of CKD in the world, led by Japan and followed by Taiwan. In Taiwan, CKD has been the eighth leading cause of death since 1997. Compared to other countries, Taiwan has a higher incidence and mortality rates, with the prevalence increasing from 1.99 % in 1996 to 9.83 % in 2003 [1], while the awareness about CKD has been low [2].

CKD causes a substantial financial burden on patients, healthcare services, and the government. Treatments of the ESRD with Renal Replacement Therapy are either expensive (haemodialysis and peritoneal dialysis) or complex (transplantation). Taiwan has about 0.1-0.2 % of the population receiving dialysis - contributing to about 7 % of the total budget of the National Health Insurance (NHI) program [3]. The association of CKD with other chronic diseases also exacerbates the situation. From the public health perspective, it is therefore imperative to be able to predict the trends in terms of CKD prevalence, so that timely decisions can be taken by the decision-makers (ministries, insurers, hospital managers, etc.) to mitigate a potential surge in the number of cases. Such mitigation measures can include enhanced population screening for CKD-related risks and awareness campaigns, as it has been demonstrated that the patient lifestyle changes (reducing body weight, improving diet, increasing physical activity, reducing alcohol consumption, avoiding smoking, early referral to nephrologists, proper use of medication and treatments to control other risk factors) are the most effective measures to combat the exacerbation of the condition with minimal associated costs [4]. Further mitigation strategies are setting up enough facilities for haemodialysis and training the personnel.

With the availability of biomedical laboratory data, the use of machine-learning techniques in healthcare for developing disease prediction models has become common. Further, methods such as deep learning and techniques like ensemble learning have greatly improved the predictive power of machine learning models. By deriving features from Electronic Health Records (EHR), accurate disease prediction models can be developed [5,6]. On a patient-level, a physician can assess the onset of CKD using laboratory tests, looking at parameters such as the glomerular filtration rate (eGFR) and with a urine test measuring the albumin-creatinine ratio, which is readily available [7]. On the other hand, from the public health perspective, laboratory data is typically not available on a large scale. However, two types of data can be readily extracted from the insurance companies' databases: diagnoses and medications for each patient's visit at the hospital.

Common approaches for developing disease prediction models with EHR data involve the collection of clinical and laboratory data from sources such as billing or claims data, discharge summaries, patient history, etc., and building models on features extracted from them. Some previous studies attempted to use longitudinal data to capture temporal patterns to develop disease prediction models for CKD. Ren Y et al. (2019) developed a predictive model for kidney disease among patients with hypertension from EHR consisting of textual and numeric information. They proposed a neural network framework based on Bidirectional long short-term memory and auto-encoders to encode the textual and numerical information, respectively. They performed under-sampling to balance the data. They achieved 89.7 % accuracy on 10-fold cross-validation [8]. Song X et al. (2020) presented a one-year prediction model using a landmark-boosting approach based on gradient boosting machines for CKD among diabetes patients with an AUROC of 0.83. They analyzed longitudinal data containing several clinical observations derived from EHR and billing data [9]. Fenglong M et al. (2018) proposed a general framework using posterior regularisation techniques that incorporate prior medical information from the EHR for prediction models. The constraint feature design in the framework took into consideration patient characteristics, underlying disease, disease duration, genetics, and family history. The patient characteristics included sex, age, and ethnicity. While using prediction models for a certain disease, the framework took into consideration the diagnosis of the underlying disease that would be related to the occurrence of the main disease to be predicted [10]. Another similar work by Katsuki T et al. (2018) predicted the risk of entering the 2nd stage diabetic nephropathy from the 1st stage using an EHR data consisting of sequences of lab test results. They used convolutional autoencoders to encode the temporal features and achieve performance better than baseline models [11].

Similarly, some studies used non-temporal EHR data to develop disease prediction models. Song X et al. (2019) extracted several significant clinical features from EHR data using an ensemble feature selection method to predict the risk of kidney disease among diabetes patients. They achieved an AUROC of 0.71 on an external validation set [12]. Jardin MJ et al. (2012) predicted kidney-related outcomes among diabetes patients using the Cox proportional hazard regression on the ADVANCE cohort, which comprises demographic, behavioral, physical information, and relevant lab values. They achieved a C statistics of 0.847 on predicting major renal events [13]. Dovgan et al. (2020) predicted the onset of renal replacement therapy three, six, and 12 months after the CKD diagnosis. For 12-months-ahead prediction, they achieved an AUC of 0.773 [14].

In this paper, we aim to develop a machine-learning model that predicts the onset of CKD within the next 6 and 12 months. The model is based on the claims data (age, sex, comorbidities, and medication) over an observation period of 24 months. Furthermore, we aim to identify which comorbidities and medications are strong predictors for the onset of CKD.

## 2. METHODOLOGY

### 2.1. Study Design

This is a retrospective study that looks at the patients who have been diagnosed with CKD and a group of patients without CKD within the chosen observation period. We define the **index date** as the time at which the patient was diagnosed with CKD (ICD code: 585, 586) for the first time. For the non-CKD group, the index date is randomly picked. We aim to predict the onset of CKD 6 and 12 months before the index data (referred to as the **lead time**). We perform the prediction by processing data two years before the lead time (referred to as the **observation time**). Figure 1 illustrates the chronology of time periods and events.

**Figure 1.**
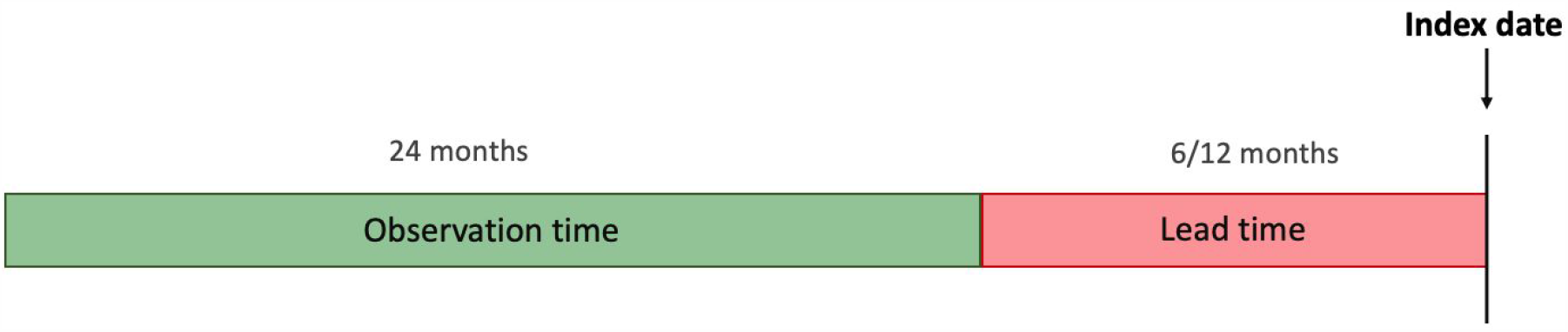
Chronology of time periods and events

### 2.2. Dataset

The study was conducted using Taiwan's National Health Insurance Research Database (NHIRD) [15], comprising patients' insurance claims data from 1997 to 2012, from which the patients' data was extracted. Each patient record consists of a patient's comorbidities or drug prescription by date. The comorbidities are represented by their ICD 9 codes and medications by their ATC codes. Figure 2 shows the distribution of patients with CKD across age and sex.

**Figure 2.**
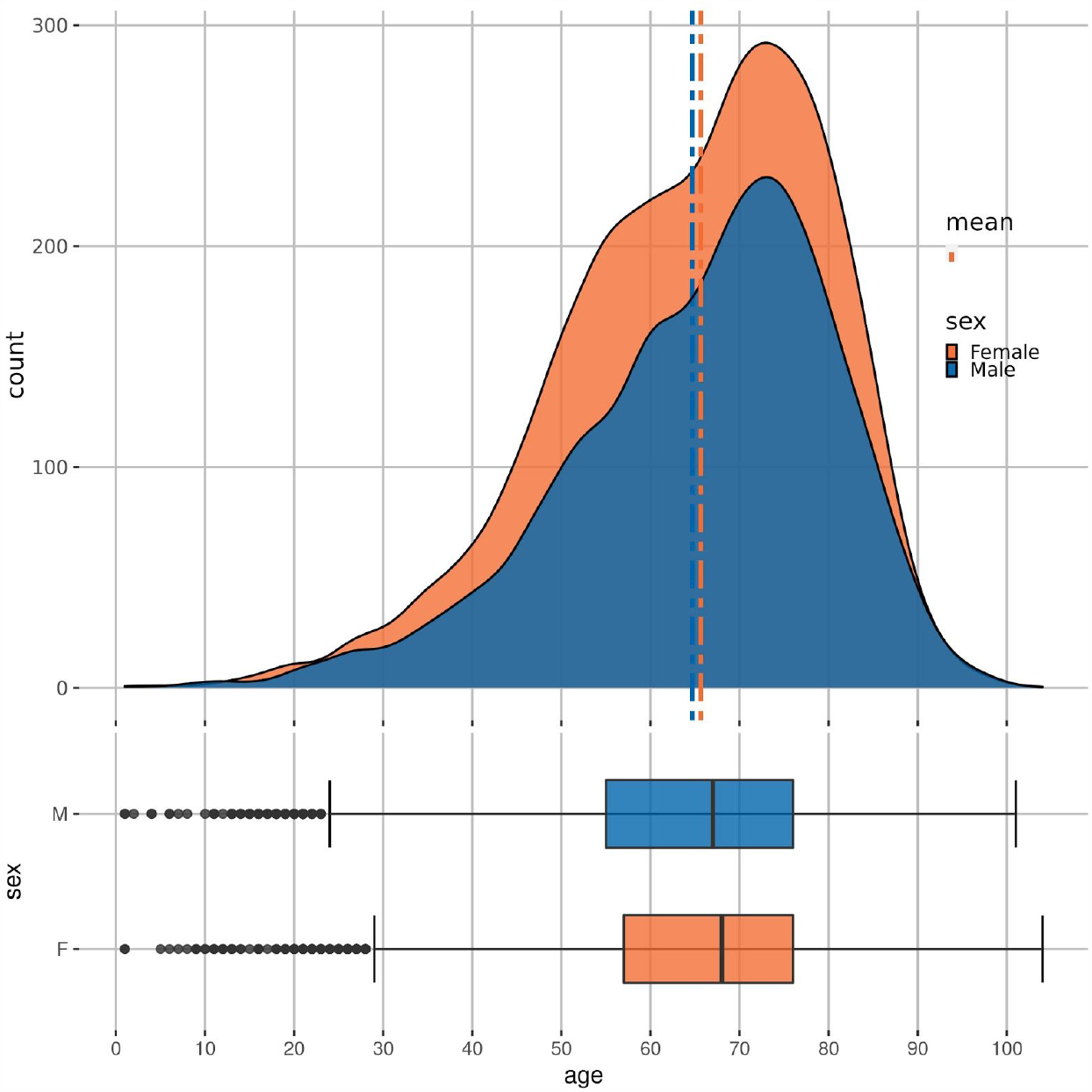
Distribution of age and sex in the CKD cohort

### 2.3. Data processing

Figure 3 shows our data processing pipeline. The first step in the pipeline consisted of data cleaning. This was done by dropping any duplicates and records with missing or incorrect values. In addition, we only included patients with age below 100.

**Figure 3.**
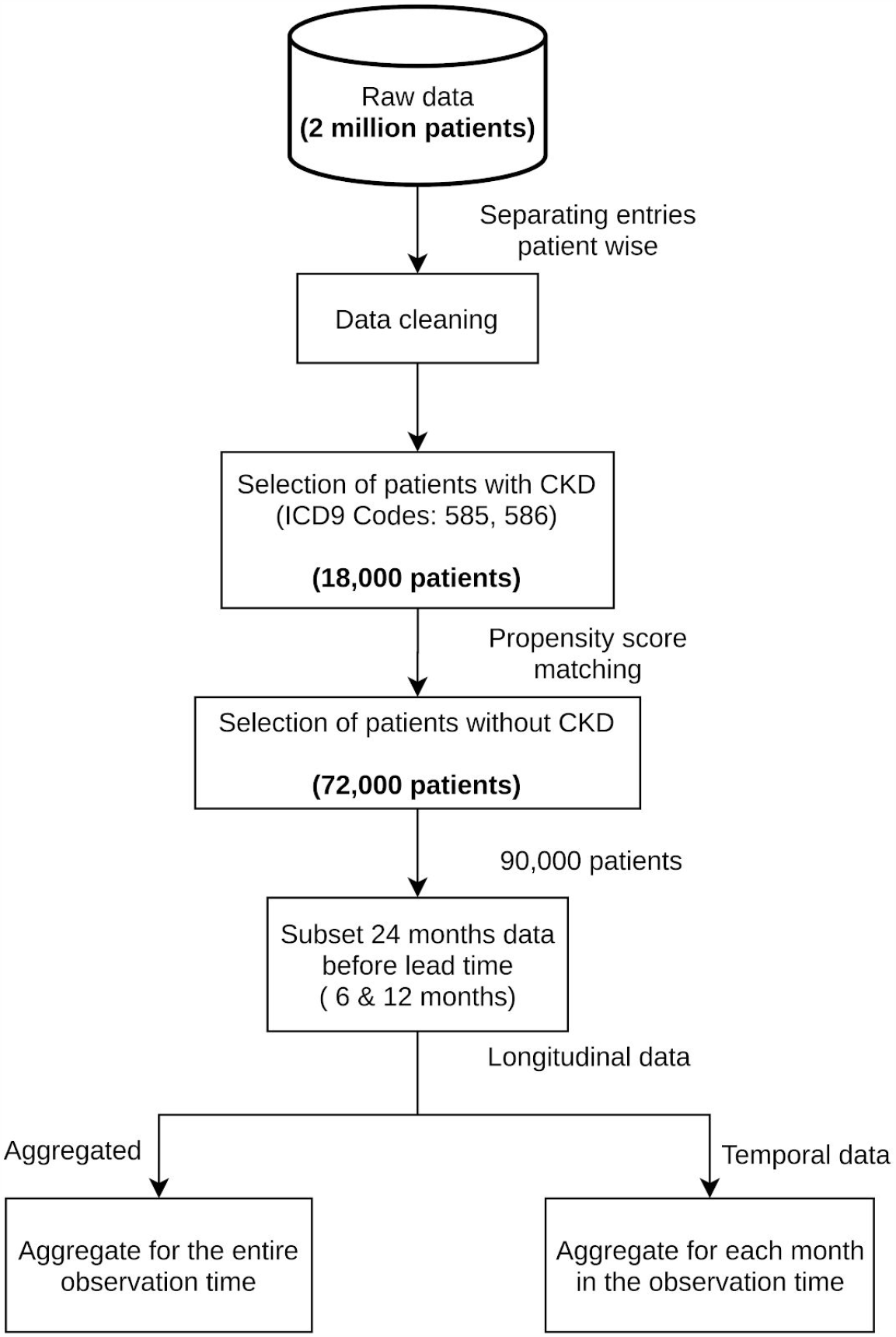
Data processing pipeline

In the second step, we selected a subsample of patients as follows. First, we selected all the patients with CKD in the database (approximately 18,000). Second, we selected 4 times as many non-CKD patients (approximately 72,000). The ratio of 1:4 was chosen as a trade-off between machine learning algorithms preferring balanced data on the one hand, and the higher number of non-CKD vs. CKD patients, on the other hand. The selection was made by using propensity score matching [16] according to age and sex, in order to obtain the same distribution of these two variables in both subsets (CKD and non-CKD). Such a selection of data reduces the selection bias and leads to better causal inferences [17]. As diseases like CKD often have a high correlation with age (see Figure 2), performing matching can lead to reliable and meaningful results.

The final step consisted of structuring the raw data into a dataset appropriate for machine learning. More precisely, we produced two datasets that included:

1. Aggregated data
2. Temporal data

To obtain the aggregated data, we discarded the temporal component by summing the occurrences of comorbidities and medications for each patient across the entire observation time. The aggregated data is thus represented with a vector where each element represents the count of comorbidities/medications throughout the observation time. Including age, sex, comorbidities, and medications, the processed data contain 1504 features.

The temporal data was obtained by aggregating the comorbidities and medications over each month during the observation time. Each patient's comorbidities and medications are thus represented with a matrix where each row represents the comorbidities (ICD codes) or medications (ATC codes), and each column represents the month of observation (i.e., months from the beginning of the lead time). The index (i, j) in the matrix represents the number of times the patient was diagnosed with/prescribed the i^th^ disease/medication during the j^th^ month from the end of the observation period.

### 2.4. Model development

We used the following modeling algorithms of different types on the preprocessed dataset from packages like Scikit-learn [18], XGBoost [19], LightGBM [20], and TensorFlow [21].

#### Logistic Regression

It is a classification algorithm that is generally used for binary classification. It uses a logistic function to find the probability of occurrence of the selected class.

#### Decision Tree

This tree-based method involves stratifying or segmenting the predictor space into simple regions. For a classification tree, each observation is predicted to the most commonly occurring class of training observations in the region to which it belongs [22].

#### Random Forest

Random Forest is an improvement of decision trees, where multiple decision trees are merged into one forest. At each split in the tree, the algorithm is not allowed to consider all the available predictors/attributes. This is implemented to avoid using the same major predictor in all of the trees, i.e., to force the trees to use lesser important predictors [22].

#### XGBoost

Extreme gradient boosting is an ensemble learning similar to Random Forest, which builds the models sequentially, applies gradient descent to decrease the error rate, and reduces overfitting with tree pruning and regularization [19].

#### AdaBoost

Adaptive boosting method is suitable for the weak base learners. It is an aggregation of many weak classifiers. The model adapts itself in every iteration sequentially based on the results of the previous iterations [23].

#### LightGBM

Light Boost is another tree-based learning algorithm. Most of the tree-based learning algorithms grow depthwise, but LightGBM grows leaf wise and, to avoid overfitting, it controls the maximum depth. This algorithm further optimizes the best split in the parallel processing, making it consume less memory and run faster [20].

#### Convolutional neural networks (CNN)

CNNs are a class of neural networks that use convolution operation on the input data to extract patterns. They are commonly used in image processing and time series analysis.

We used a 1D Conv layer, which can perform convolution across a temporal dimension. Figure 4 shows our CNN architecture.

**Figure 4.**
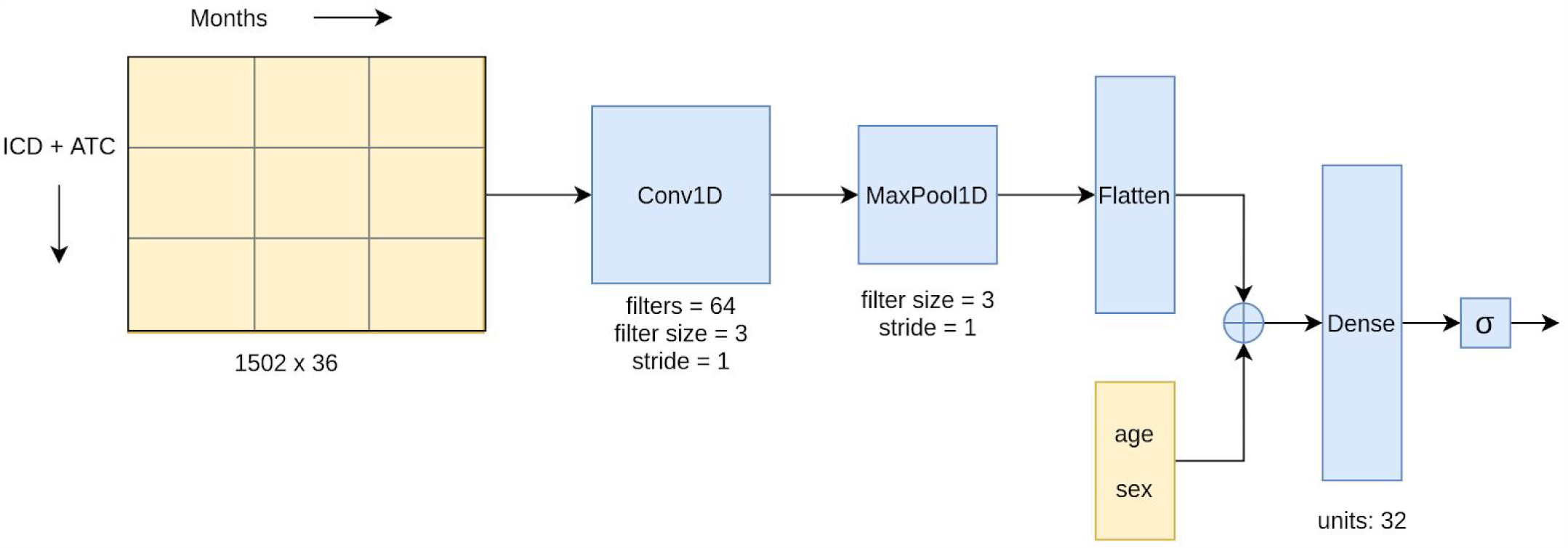
The CNN architecture used in this study

#### Bi-directional Long Short Term memory (BLSTM)

LSTMs are recurrent neural networks capable of extracting features in the forward and backward direction from a sequential input. An LSTM unit captures long-term effects, making it useful in time-series analysis.

Among the methods listed above, we used the CNN and BSLTM algorithms to model the temporal data and the remainder of the algorithms for the aggregated data. Deep learning (CNN and BLSTM) was the feasible choice to model the longitudinal data owing to its ability to handle large data inputs through GPU parallelization, while the number of features in temporal data was too large for the other algorithms. Conversely, the aggregate data was not suitable for the deep learning approaches since they are explicitly designed for two-dimensional or temporal data.

Since we do not want the predictive models to favor the non-CKD class over the CKD class due to data imbalance, we added the weights to the classes: 4 to the minority class (CKD) and 1 to the majority class (non-CKD). However, since all the methods are not able to handle class weights, we also used the following approaches. For the CNN algorithm, we assigned weights of 1 and 4 to the loss for CKD and non-CKD classes, respectively. For the AdaBoost and gradient boosting algorithms, we oversampled the minority class as the Scikit-learn library does not provide an interface to assign class weights for these algorithms.

The Logistic Regression required a further data preprocessing step, namely, data normalization. This was achieved by scaling the age by the maximum age and binarizing the diagnosis and drug prescription counts. The binarization was done by setting any non-zero values to 1.

Some of the models require parameter tuning. A subset of methods' hyperparameters was tuned with 5-fold cross-validation on the training set. The test set on which the final models were evaluated was not used for tuning, to obtain an unbiased evaluation of the models. In some cases, using the default parameters was already sufficient. The values of the hyperparameters used in the final models are listed in the Supplementary Information.

### 3. RESULTS

### 3.1. Evaluation metric

We split the whole dataset into an 80 % training set and a 20 % test set. We train our models on the training set (the same one which was used for hyperparameter tuning with 5-fold cross-validation) and report their performance on the test set. The train-test and cross-validation splits are performed by stratifying on the target label. We primarily use the area under the ROC curve (AUROC) as our evaluation metric [24]. The Receiver Operating Characteristic (ROC) curve, which is a plot of the true positive rate (Recall, Sensitivity) against the false positive rate (1 - Specificity) at different probability thresholds, assesses the discriminative ability of a given binary classification model. It illustrates the model's ability to identify positive instances while minimizing false alarms correctly. The plots in Figure 5 show the ROC curve for different algorithms on 6- and 12-month data.

**Figure 5.**
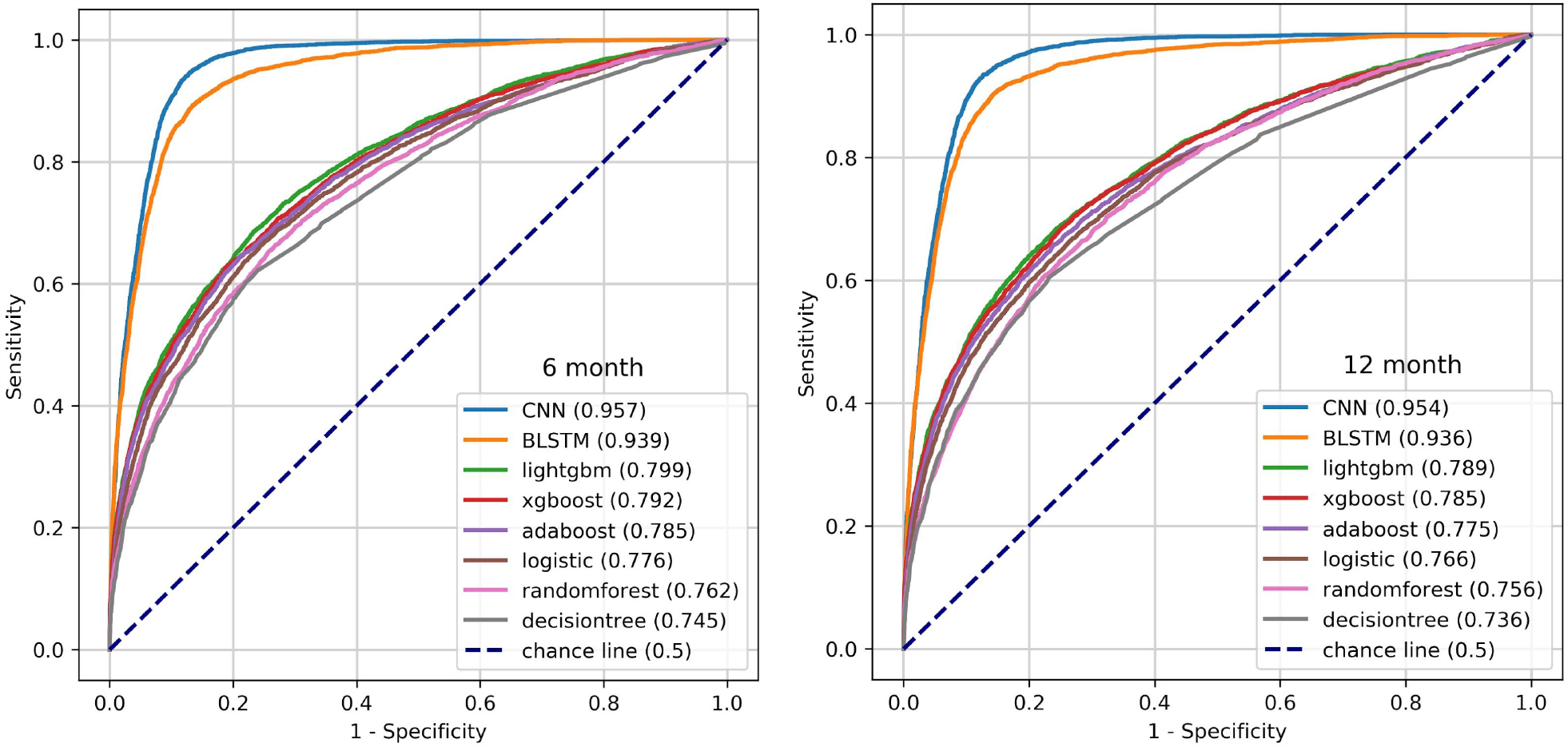
ROC curves for different models

The confusion matrices for the CNN models are shown in Table 1, where we can observe that the models miss few positive instances and raise only a modest number of false alarms considering the larger number of negative instances.

**Table 1.** Confusion matrices of the CNN model, showing the number of instances in each class, while the fraction of predicted instances is in parentheses

**12-month model**
|  | Predicted<br>**Negative** | Predicted<br>**Positive** |
| --- | --- | --- |
| Actual<br>**Negative** | 12841<br>(0.9) | 1359<br>(0.1) |
| Actual<br>**Positive** | 423<br>(0.12) | 3169<br>(0.88) |

**6-month model**
|  | Predicted<br>**Negative** | Predicted<br>**Positive** |
| --- | --- | --- |
| Actual<br>**Negative** | 13035<br>(0.91) | 1288<br>(0.09) |
| Actual<br>**Positive** | 411<br>(0.11) | 3186<br>(0.89) |

**Table 2.** Performance metrics for 6-month data

| Algorithm | Accuracy | F1 | Precision | Recall or<br>Sensitivity | Specificity | AUROC |
| --- | --- | --- | --- | --- | --- | --- |
| CNN | **0.905** | **0.789** | **0.712** | **0.886** | **0.91** | **0.957** |
| BLSTM | 0.873 | 0.737 | 0.63 | 0.888 | 0.869 | 0.939 |
| lightgbm | 0.759 | 0.526 | 0.435 | 0.664 | 0.782 | 0.799 |
| xgboost | 0.777 | 0.527 | 0.46 | 0.619 | 0.817 | 0.792 |
| adaboost | 0.79 | 0.522 | 0.483 | 0.569 | 0.846 | 0.785 |
| logistic | 0.744 | 0.505 | 0.414 | 0.648 | 0.768 | 0.776 |
| randomforest | 0.788 | 0.484 | 0.475 | 0.493 | 0.862 | 0.762 |
| decisiontree | 0.744 | 0.486 | 0.408 | 0.601 | 0.78 | 0.745 |

**Table 3.** Performance metrics for 12-months data

| Algorithm | Accuracy | F1 | Precision | Recall or Sensitivity | Specificity | AUROC |
| --- | --- | --- | --- | --- | --- | --- |
| CNN | **0.9** | **0.781** | **0.7** | **0.882** | **0.904** | **0.954** |
| BLSTM | 0.883 | 0.748 | 0.66 | 0.863 | 0.888 | 0.936 |
| lightgbm | 0.749 | 0.519 | 0.424 | 0.671 | 0.769 | 0.789 |
| xgboost | 0.764 | 0.518 | 0.442 | 0.626 | 0.799 | 0.785 |
| adaboost | 0.786 | 0.516 | 0.475 | 0.564 | 0.842 | 0.775 |
| logistic | 0.73 | 0.492 | 0.397 | 0.647 | 0.751 | 0.766 |
| randomforest | 0.782 | 0.476 | 0.464 | 0.488 | 0.857 | 0.756 |
| decisiontree | 0.736 | 0.479 | 0.399 | 0.601 | 0.77 | 0.736 |

We also report accuracy, precision, recall, F1 score, and sensitivity on the CKD class for the models. Tables X and Y summarise the performance of the models on 6- and 12-month data.

To understand the amount of training data required to build a CNN model, we trained multiple CNN models by varying the size of the input training data and recorded the AUROC performance on our test set. The plot in Figure 6 presents the results, which indicate diminishing returns of additional instances after 20,000

**Figure 6.**
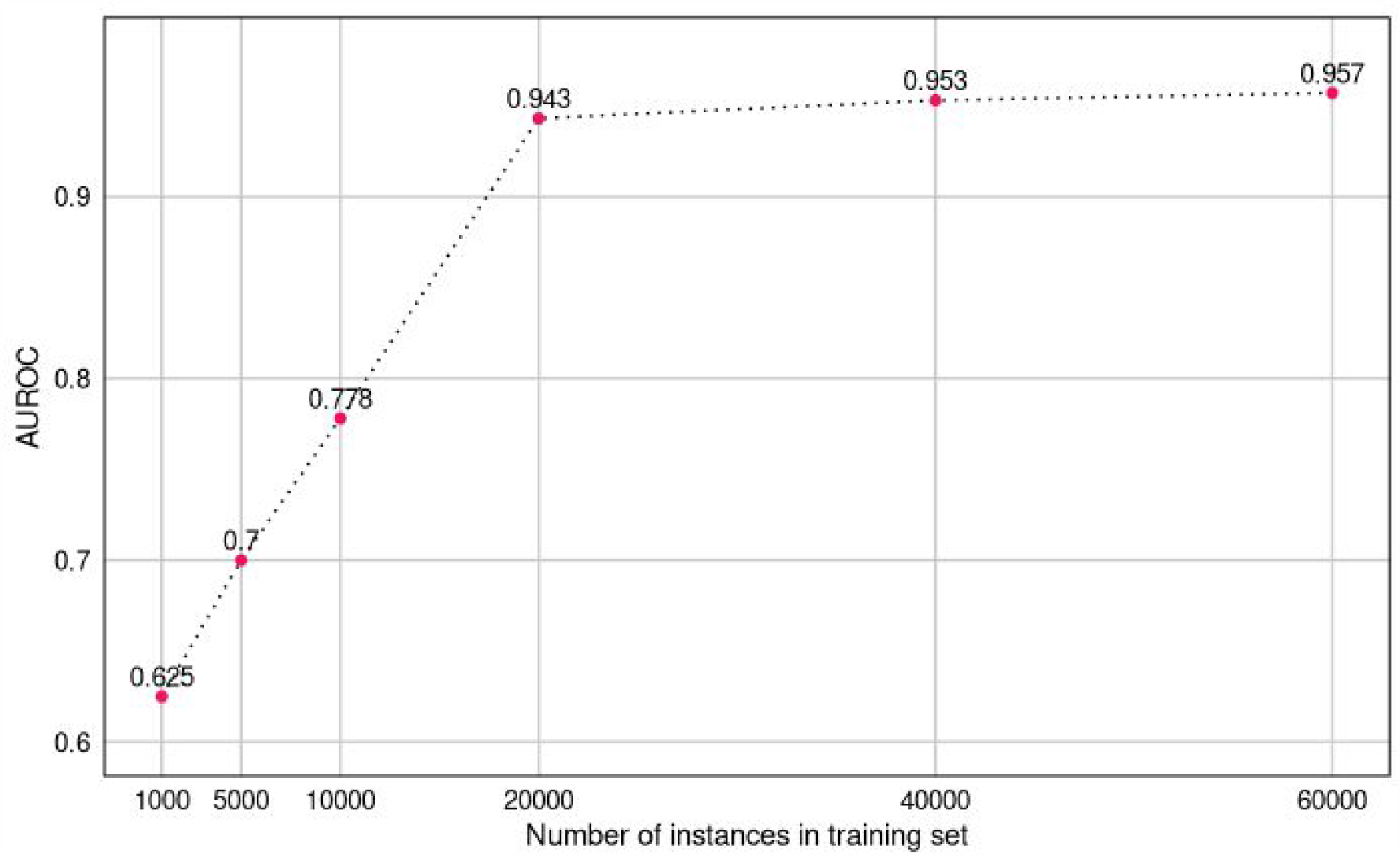
Performance of the CNN model across different size of training data

### 3.2. Feature importance from boosting methods

One of the advantages of using tree-based algorithms is that they are interpretable (unlike deep learning algorithms, which have complex feature interactions and a large number of model parameters, so they act as a “black box”). While a human can simply look at plain decision trees, interpretation is more difficult for boosting and forest models. However, feature importance can still be derived from such models using the computed information gain values.

The information gain measures the reduction of the entropy when dividing a set of data into subsets, where entropy is a measure of uncertainty. For example, if in a set of patients, the probability of having CKD is 50%, the entropy is the highest. If we can divide this set into pure sets containing only CKD or only non-CKD patients, the resulting entropy is 0. For each attribute, we can measure its contribution to the reduction of entropy, *i.e*., its contribution to the improved explanation of the target variable.

We plot the feature importance for men and women to identify the key predictors associated with sex. The normalized feature importance was derived from lightgbm models.

The results unveiled in Figure 7 highlight the factors that influence the development of CKD, and are observed 6 and 12 months prior to diagnosis of CKD. Diabetic nephropathy is a common complication of Diabetes mellitus (DM). In addition, medications related to DM, gout and hypertension are seen as the risk factors. Due to kidney disease filtration of uric acid is compromised thus causing gout. However, gout may also lead to the progression of kidney disease.

**Figure 7.**
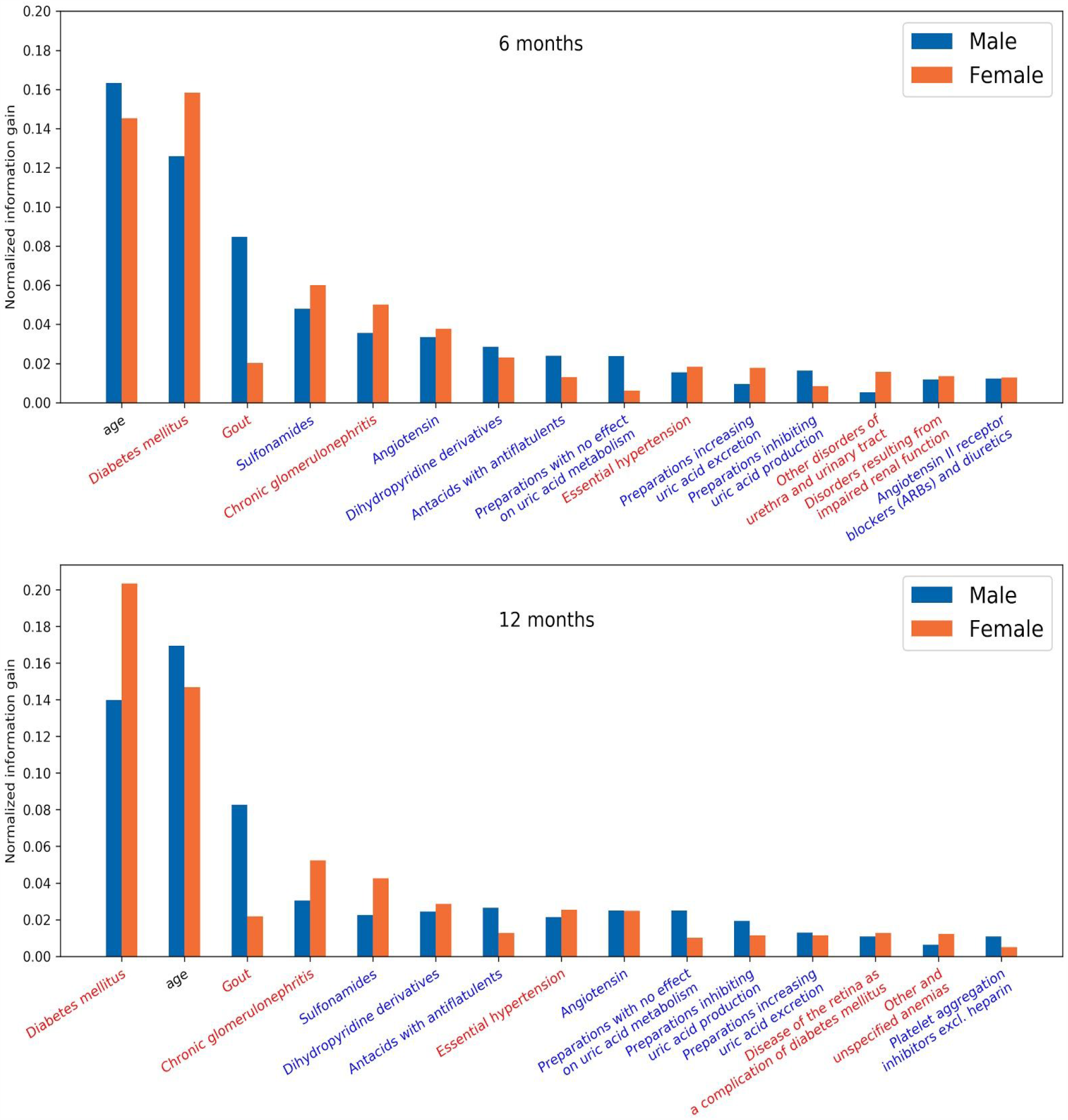
Feature importance for the LightGBM models for men and women for 6 and 12 months. The color of the x-axis labels is related to the type of feature: comorbidities are red, medications are blue, and age is black.

## 4. DISCUSSION

Among the machine-learning models, deep neural networks (CNN and BLSTM) outperformed the classical models considerably. This is interesting as they are not as well established in health risk prediction as in some other fields (most notably computer vision). Considering that our networks were not very large or complex, the most likely reason seems to be that they took advantage of temporal information, which was not available to the classical models. Trying to provide such temporal information to the classical machine-learning algorithms resulted in too many features, though, and addressing this issue with feature selection and dimensionality reduction did not prove fruitful, either, suggesting that the redundancy in the features (comorbidities and medications) is modest.

In contrast to the commonly used approach that processes laboratory data in addition to other patient data, we developed a method that does not require laboratory data, but processes only patients' diagnoses, prescriptions and basic demographic data (i.e., age and sex), since such data is typically available on a larger scale.

Looking at the most prominent features in the LightGBM models, we see that age is among the strongest predictors in both 6 and 12 models. Among comorbidities, diabetes mellitus is the strongest predictor in both models, followed by gout, chronic glomerulonephritis, and essential hypertension. The medications most associated with the model are sulfonamides, followed by angiotensin, dihydropyridine derivatives, and antacids with antiflatulents. There are some differences in feature importance for men and women. While age appears to be a slightly stronger predictor for men and diabetes mellitus for women, the most obvious difference is in gout, where the feature importance is much stronger for men than for women.

It is reasonable to see age as a strong predictor as CKD as the condition typically appears in older people. Diabetes mellitus and gout are both strongly connected with a decreasing kidney function, as are chronic glomerulonephritis and essential hypertension. Gout is a stronger feature in men, which is probably related to different diets and lifestyles [25]. Sulfonamides are a group of antibiotics. They are often used to treat urinary tract infections, and repeated infections may lead to the onset of CKD. Kidney damage is one of the severe complications of sulfonamide therapy. Dihydropyridines are vasodilators and are used to treat hypertension. Angiotensin is also used for blood pressure regulation. Medications dealing with uric acid production and secretion are relevant since they are used to treat gout. We suspect that antacids with antiflatulents are not directly connected with the onset of CKD. These medications are usually prescribed to people with several other comorbidities as one in a series of medications, to reduce the burden of other medications on the gastrointestinal tract. This is clearly correlated with age, as older people tend to have more conditions that require polymedications.

## 5. LIMITATIONS

Our study was built upon the data extracted from Taiwan NHI that includes age, sex, comorbidities, and medications. However, laboratory test results are not included. Therefore, our approach is appropriate for a population study and not to assist the clinicians in assessing the risks for an individual patient. For clinical practice, the decisions based on laboratory test results are far superior.

Furthermore, the fact that the study was carried out on the data from Taiwan, thus limited by geography, imposes constraints on the generalizability of the model to the global population or a different region. The presence of noise in the data from human and technical errors, which are difficult to identify, may affect the performance of the model to some degree as well.

## 6. CONCLUSION

In this study, we developed and evaluated a series of artificial intelligence-based models considering minimum variables such as sex, age, comorbidities, and medications. These models predict patients' risk of developing chronic kidney disease within the next 6 or 12 months. Among various models tested, convolutional neural networks (CNN) performed best, with the AUROC metric of 0.957 and 0.954 for 6 and 12 months, respectively. To see which features are the most prominent for prediction, we looked at the tree-based LightGBM model. The most prominent features included diabetes mellitus, age, gout, and use of sulfonamides and angiotensins, which are all reasonable in view of CKD.

From the policy-maker's point of view, these ML-based models can be efficiently used in resource management and initiating public health initiatives such as closely monitoring and early detection of CKD. Clearly, for application of such models into clinical practice dealing with individual patients, the feature set would have to be expanded to include laboratory measurements and possibly lifestyle information which falls within the scope of future work.

## Data Availability

The data that support the findings of this study are available on request. The data has been taken from Taiwan's National Health Insurance Research Database (NHIRD).

## ACKNOWLEDGMENTS

The authors acknowledge the financial support from the Slovenian Research Agency (research core funding No. P2-0209). This work is part of the CrowdHEALTH project that has received funding from the European Union's Horizon 2020 research and innovation program under grant agreement no. 727560 (JSI) and the Ministry of Science and Technology under project no. 106-3805-018-110 (TMU).

